# Effects of post tuberculosis lung disease on survival in HIV-infected individuals with pulmonary hypertension: Insights from the Pan African Pulmonary Hypertension Cohort (PAPUCO) study

**DOI:** 10.1101/2023.08.20.23294338

**Authors:** Patrick D.M.C. Katoto, Sandra L. Mukasa, Karen H. Wolmarans, Mahmoud U. Sani, Kamilu M. Karaye, Irina Mbanze, Albertino Damasceno, Ana O. Mocumbi, Anastase Dzudie, Karen Sliwa, Friedrich Thienemann

## Abstract

**Introduction:** Post-tuberculosis lung disease (PTLD) bears high mortality rates, primarily attributed to pulmonary vascular and cardiovascular complications. We investigated the impact of tuberculosis (TB) history on pulmonary hypertension (PH) prognosis within an HIV-burdened region.

**Methods:** We acquired sociodemographic, clinical, and echocardiographic data on 206 PH adults from the Pan African Pulmonary Hypertension cohort (PAPUCO), a prospective cohort study undertaken in four African countries. Cox-hazard regression models were constructed to assess how TB history interacts with diabetes, HIV-infection, and other chronic lung diseases (CLD), impacting death risks in PH patients.

**Results:** Among the participants, a history of TB, diabetes, and other CLD was found in 23%, 8%, and 12% respectively. Of the 47 (35%)/134 participants living with HIV-infection, 62% exhibited HIV/TB coinfection, with 45% experiencing recurrent TB episodes. Individuals with TB history faced a 1.82-fold higher PH-related mortality risk (adjusted Hazard Ratio [aHR]: 1.84; 95%CI: 1.00, 3.39; p=0.049). Concurrent TB and comorbidities amplified death risks for PH patients, significantly affecting CLD (aHR: 3.10; 95%CI: 1.47, 6.53; p=0.003), and showing borderline impact for HIV co-infection (aHR: 2.10; 95%CI: 0.97, 4.54; p=0.059), while not significantly influenced by diabetes history (aHR: 2.39; 95%CI: 0.32, 18.00; p=0.4), although clinically relevant.

**Conclusion:** Nearly one in every four patients diagnosed with PH in Africa have a history of TB and one in every three have HIV infection, which dramatically reduces their odds of survival. Our findings constitute a call to action to effectively address the neglected burden of PH among millions of patients suffering with TB diseases.

## Introduction

Prior to the coronavirus (COVID-19) pandemic, tuberculosis (TB) was the leading cause of death from a single infectious agent, surpassing HIV/AIDS. A 6-month drug regimen can successfully treat about 85 percent of people who develop TB disease. Multisectoral action to address TB determinants such as poverty, undernutrition, HIV infection, smoking, and diabetes can also reduce the number of people becoming infected and developing disease (and thus the number of deaths caused by TB). Some countries have already reduced their TB disease burden to less than 10 cases and 1 death per 100,000 people per year^1^.

Many people who survive TB face ongoing disability and an increased risk of death^2^. The impact of post-TB sequelae, on the other hand, is frequently overlooked in policy analyses and disease burden estimates. Since 2000, an estimated 58 million people have survived TB, but many will develop post-tuberculosis lung disease (PTLD). PTLD is caused by a complex interaction of organism, host, and environmental factors, and has long-term consequences for respiratory and cardiovascular health. PTLD is a group of disorders that affect the large and small airways (bronchiectasis and obstructive lung disease), lung parenchyma, pulmonary vasculature, and pleura, and can be complicated by co-infection and haemoptysis^3^. Multiple TB episodes, drug-resistant TB, delays in diagnosis, and possibly smoking are all risk factors for PTLD. Diabetes and HIV infection, for example, raise the risk of TB by 1.5 (1.3-1.8) and 18 (15-21), respectively^1,3^. When compared to the acute phase alone, DALYs associated with post-TB disease increase the total burden of TB disease by 91%. The DALYs associated with PTLD are affected by differences in survival: some countries with a high TB burden had post-TB DALYs greater than 20 DALYs per 1000 people, compared to the global average of 75 DALYs per 1000 people. Factors associated with higher DALYs per incident case include young age, HIV infection, living in a high-incidence country, and not receiving TB treatment ^4,5^.

TB and cardiorespiratory diseases are two examples of communicable and noncommunicable diseases that occur in the same patient population, albeit in a different patient population^6^. TB is associated with pulmonary hypertension (PH), which is associated with a poor prognosis^7^. PH, is a complex disease and is classified into five groups **-** group 1 pulmonary arterial hypertension (PAH), group 2 PH associated with left heart disease (PH-LHD), group 3 PH associated with lung disease (LH-LD), group 4 PH associated with pulmonary artery obstruction, most commonly chronic thromboembolic pulmonary hypertension (CTEPH), group 5 PH with unclear and/or multifactorial mechanisms^8^. Due to both human and material challenges, PH remains an understudied topic in low- and middle-income countries (LMICs) and particularly in PTLD, even though approximately 80% of the estimated 1 million people with PH live in LMICs. Because right heart catheterization is such a technically demanding diagnostic procedure, millions of TB survivors at risk of PH are unlikely to be identified. As a result, many will go untreated, exacerbating TB-related morbidity. Management decisions appear to be largely influenced by anecdotes in the absence of sufficient data. Patients with history of TB are more likely to be categorized in group 3, which has six subgroups (obstructive lung disease or emphysema, restrictive lung disease, lung disease with mixed restrictive/obstructive pattern, hypoventilation syndromes, hypoxia without lung disease and developmental lung disorders) and consequently their treatment algorithms are extrapolated from this class^4,8–11^.

In this secondary analysis study of the Pan African Pulmonary Hypertension Cohort, a unique contemporary multi-country registry available to the continent^12–15^, we aimed to assess the effect of TB history on the prevalence and hazards of death associated with PH in the region with an important burden of HIV/TB coinfection, while also considering other emerging TB risk factors such as diabetes and chronic lung diseases on the rise on the continent. Our findings should rekindle debate about the proper management of noncommunicable diseases associated with TB care, as well as the need for effective universal health care to address the intersection of chronic communicable diseases and NCDs in Africa and other regions.

## Methods

### Ethical approval

Each participating institution had prior approval from their respective institutional ethics review boards. This research was conducted in accordance with the principles outlined in the Helsinki Declaration. All participants gave their informed consent in writing before being enrolled in the study, and HIV testing was performed following all applicable ethical and regulatory guidelines.

### Cohort profile

The PAPUCO registry is the largest ongoing cohort study of PH in Africa; its overall design, goals, and specific methods have all been previously described in detail^12^ as well as pre-registered at ClinicalTrials.gov (NCT02265887). The STROBE guidelines for reporting observational outcomes have been followed as closely as possible in the registry^16^. In 2011, the PAPUCO research group began collecting data to instigate a prospective cohort study of de novo PH cases that would be representative of the entire African continent. As a result, the registry includes data from nine facilities providing specialist care referrals in Cameroon, Mozambique, Nigeria, and South Africa. To participate, patients had to be newly diagnosed with PH using standardized clinical and echocardiography criteria, able to return for 6-month follow-up if alive, and older than 18 years. Eligible centers had to have the following characteristics: the ability to perform echocardiography, expertise in the diagnosis of PH according to the World Health Organization classification, expertise in the clinical management of patients with right heart failure (RHF), and the ability to review patients at 6-month follow-up. Following the ESC and European Respiratory Society’s (ERS) guidelines for the diagnosis of PH, a diagnostic algorithm was established to detect PH and PH-LHD in resource-constrained settings without access to right heart catheterization^17, 18^. Specialist cardiologists diagnosed PH based on the presence of dyspnea, fatigue, peripheral oedema, and other cardiovascular symptoms, ECG, and chest X-ray changes consistent with PH, and a transthoracic echocardiogram showing an elevated right ventricular systolic pressure (RVSP) of 35 mm Hg and above in the absence of pulmonary stenosis and acute RHF. CT scans, ventilation/perfusion scans, and right heart catheterizations were performed if deemed necessary by the attending physician. For this study, we used the PAPUCO dataset, which includes information on history of both TB and HIV-infection.

### Variables of interest

Firstly, we recorded patient’s demographic and ethnic background, medical history and information on all major cardiovascular diagnoses and up to five non-cardiovascular diagnoses according to International Classification of Diseases (ICD) 10 coding. Secondly, we assessed patient’s clinical status after a comprehensive physical examination and symptom scoring. The World Health Organization (WHO) Functional Class (FC), 6-minute walk test (6-MWT), and Karnofsky performance score were all included in the sequence of functional tests. Thirdly, each participant had a chest x-ray, 12-lead electrocardiogram (ECG), and echocardiography. Fourthly, we kept a record of every prescription therapy as well as the related supportive care that was obtained. Finally, we collected patient clinical outcomes such as hospitalization and mortality during scheduled 6- and 12-month follow-ups. At this point of time, a verbal autopsy was performed to record survival. Each case was evaluated by at least two investigators to confirm that the data was complete and authentic. All study data were collected and stored in a secure central database utilizing electronic case report forms (web-based platform).

### Statistical analysis

We collected survival data from the PAPUCO registry using a Microsoft Excel sheet for 206 individuals with a complete record of a TB history. R (The R Foundation for Statistical Computing, Vienna, Austria) V4.0.5 and GraphPad prism (v8.01) were used for data cleaning and analysis. Patients were divided into two groups based on whether they had a history of TB or not, and whether they were HIV-infected or not. Where applicable, continuous data were provided as mean and standard deviation [SD], and categorical data as frequencies (percent). We utilized the chi-square test to compare proportions between two groups, and the t-test/ Fisher’s exact test to compare two means. Given the endemicity of TB in the WHO area of Africa, we constructed Kaplan-Meier survival estimates and used the log-rank test to analyze the influence of recurrent TB on PH patient survival. The impact of TB history on the risk of mortality in PH patients was studied using multivariable Cox proportional hazards regression models. To examine the proportional hazard assumption, the Nelson-Aalen cumulative hazard function and the Schoenfeld residuals test were utilized. Four multivariable models were developed, one of which took into account TB history in isolation from other relevant comorbidities, and the other three of which looked at the interaction between TB and diabetes, HIV status, and other chronic lung illnesses simulating real life possibilities. All models were adjusted for age, gender, and RVSP. We used the hazards ratio and its associated 95%CI to characterize the magnitude of the association. All p-values were two-sided, with p 0.05 being the significance level.

## Results

### Characteristics of 206 participants with PH in the PAPUCO registry by TB history

Of the 206 participants with mean age of 50 years, 47 (23%) had TB, 17 (8%) had diabetes, and 24 (12%) had other chronic lung diseases (**Table 1**). Patients with a history of TB who developed PH were much younger (39 vs 53 years), lived in temporary housing, and were mostly unemployed. Furthermore, they had a history of tobacco smoking (current or previous smoking), rheumatic heart disease, pulmonary embolism, and chronic lung diseases, but not of schistosomiasis or hypertension. Patients with PH and a history of TB complained less about fatigue (77 % vs 92 %) than those without a history of TB. They also performed well during the six-minute walk test, either for the total distance walked in six minutes (296-meter vs 217-meter) or for the proportion who walked less than 300 m in six minutes (44 % vs 68 %). Similarly, their BMI and systolic pressure were lower than those without a TB history, and they had raised JVP and S3 gallop. We also found that PH patients with a history of TB had higher platelet levels, lower creatinine levels, and higher CRP levels than PH patients without. While PH-targeted therapy was highly uncommon among study participants (12%), PH patients with a history of TB had a lower risk of receiving phosphodiesterase treatment than those without such a history (6 % vs 0 %).

**Table 1:**
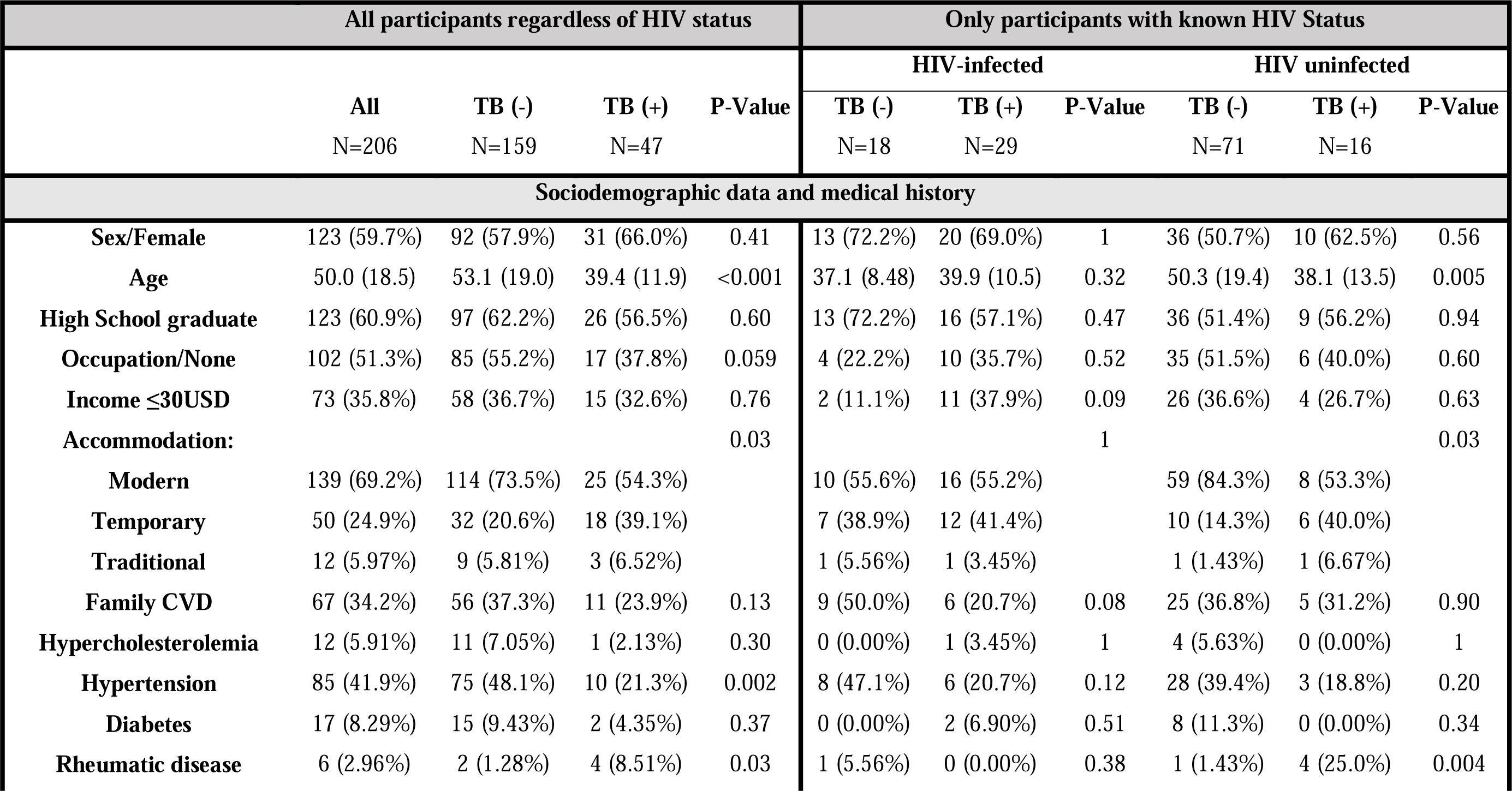
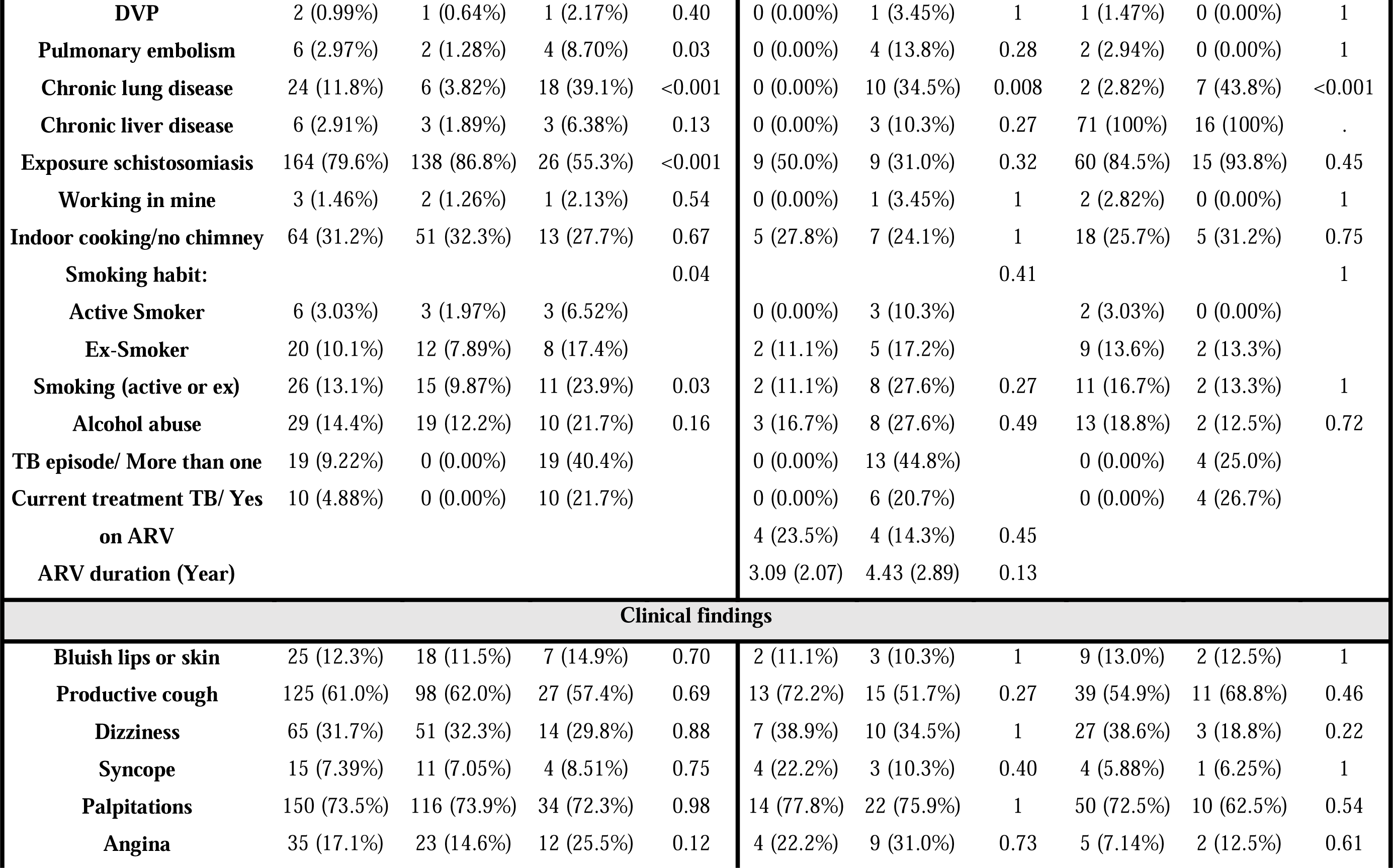
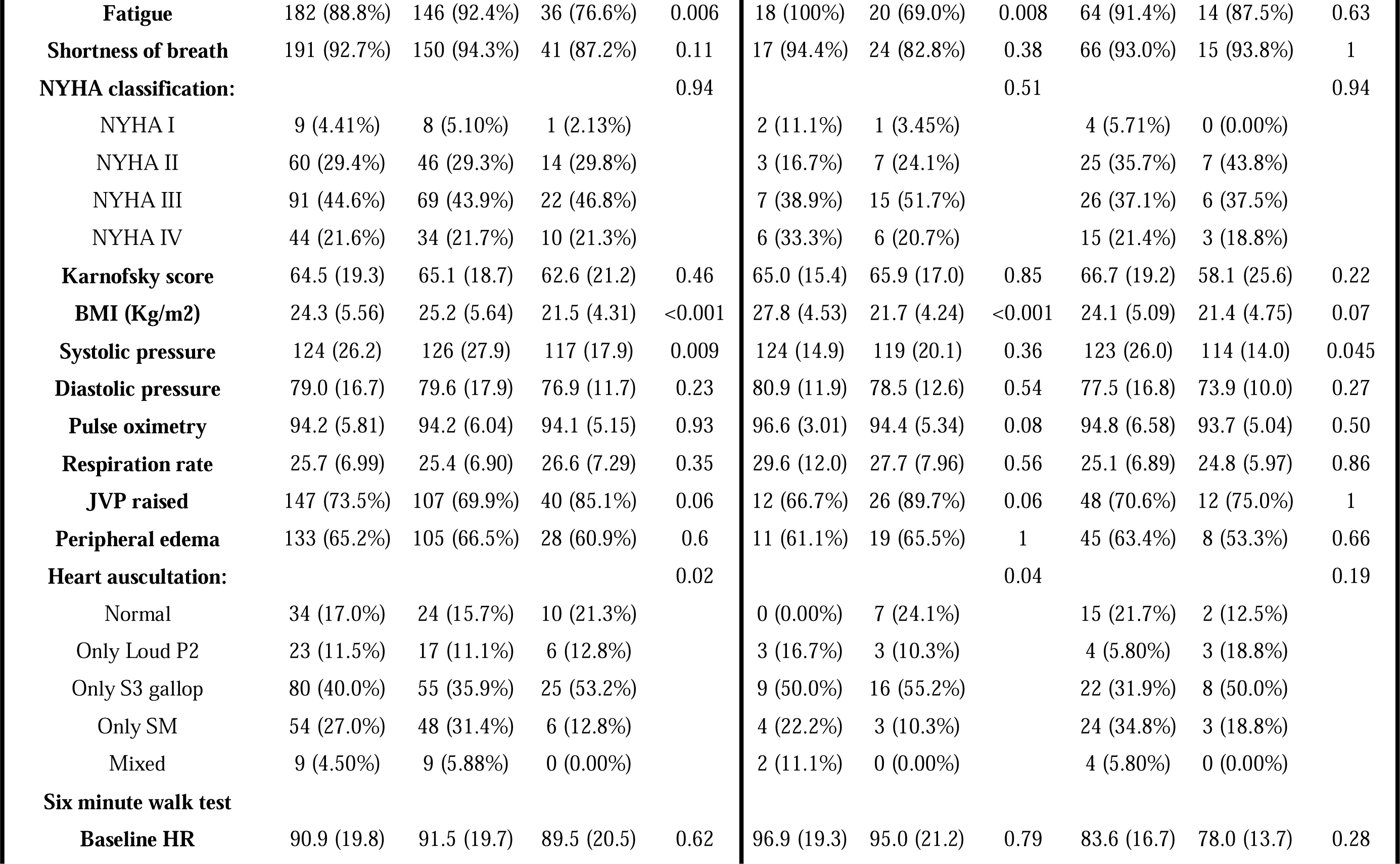
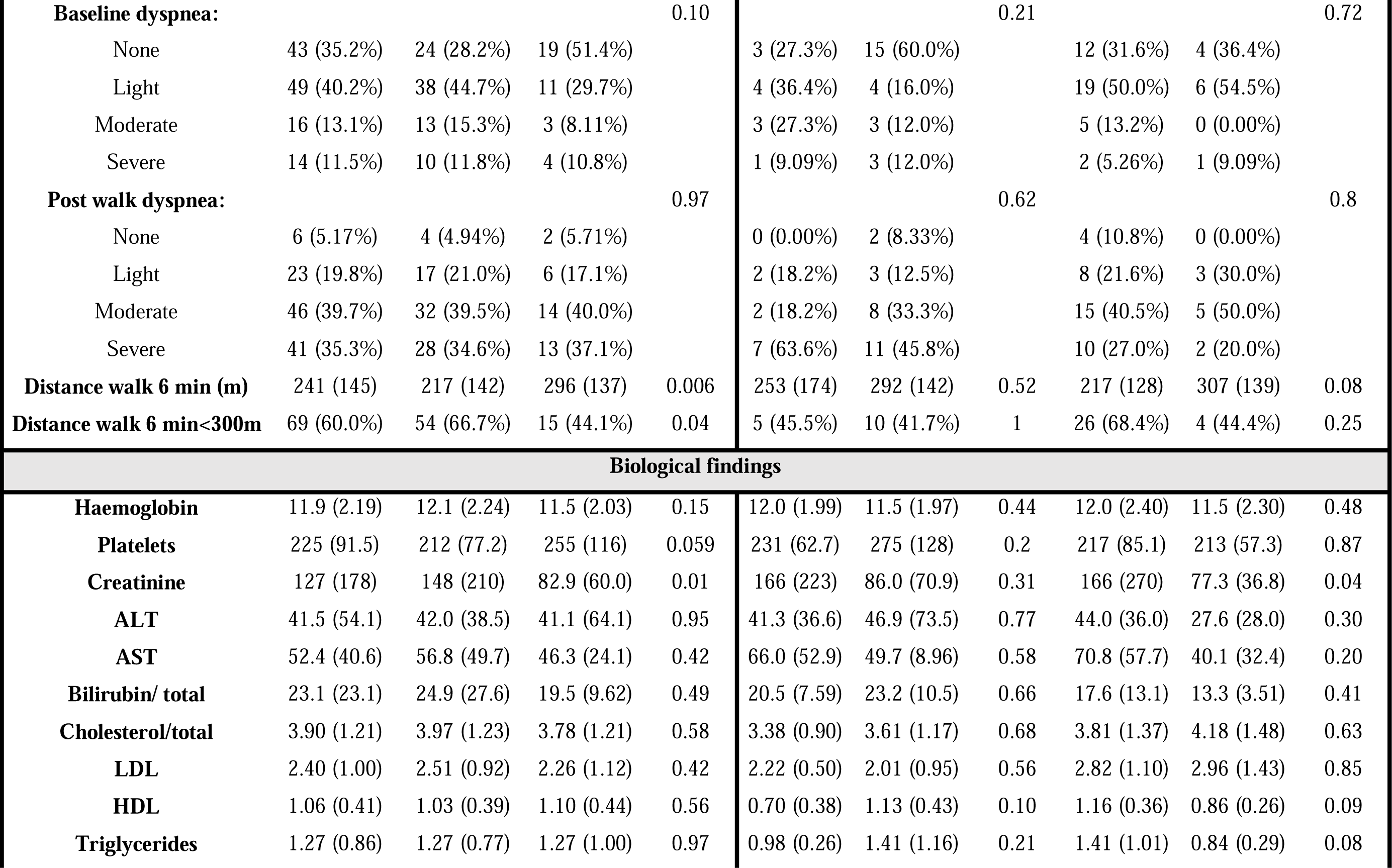
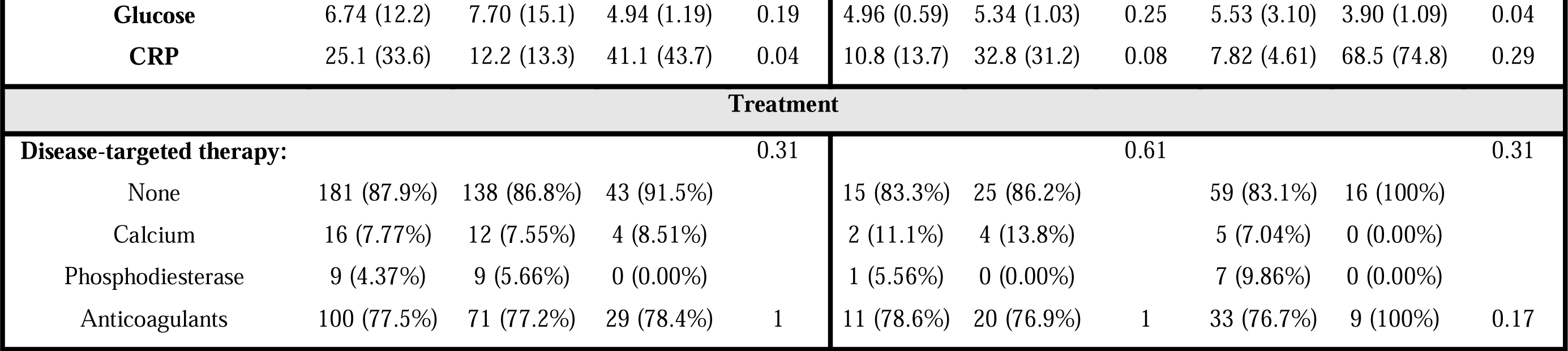
Sociodemographic, clinical, biological and treatment characteristics of 206 patients included in the Pan African Pulmonary Hypertension Cohort by HIV-infection status and by history of tuberculosis.

**Table 2** summarizes the findings of the ECG, chest X-ray, and echocardiography. P pulmonale (37 % vs 11 %) and RV hypertrophy strain (35 % vs 16 %) but not LV hypertrophy (12 % vs 30 %) and atrial fibrillation (2 % vs 20 %) were more common in PH patients with a TB history than in those without a TB history. According to echocardiographic data, PH patients with a TB history had considerably lower RVSP (58 vs 64 mmHg) and higher fractional shortening (31 vs 26%) than those without a history of TB. They were more likely to have PH-LD (36 vs 4%) than their counterparts, who were also more likely to have PH-LHD (78 vs 36 %).

**Table 2:**
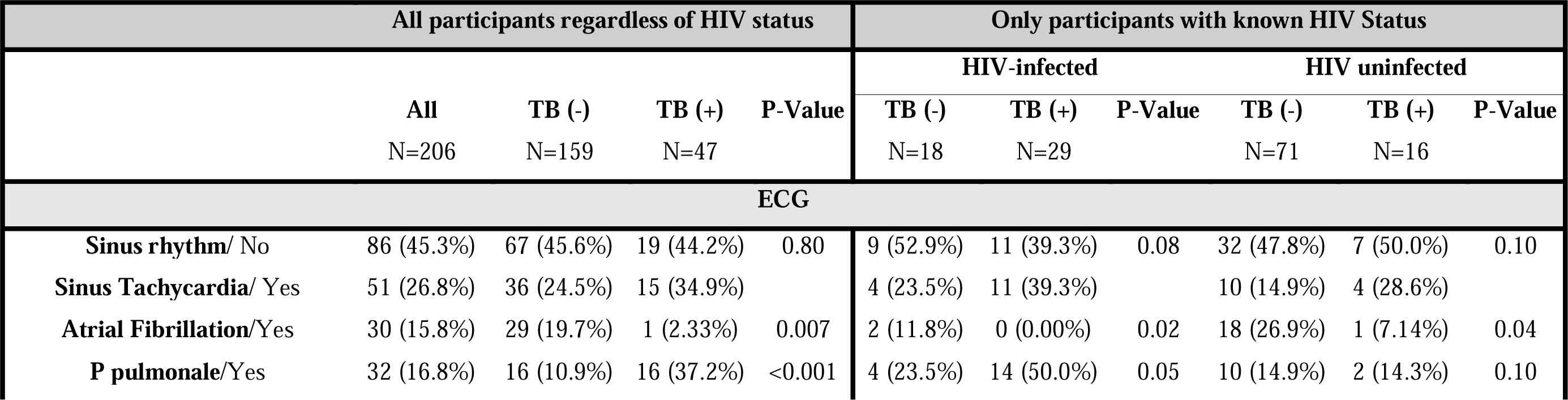
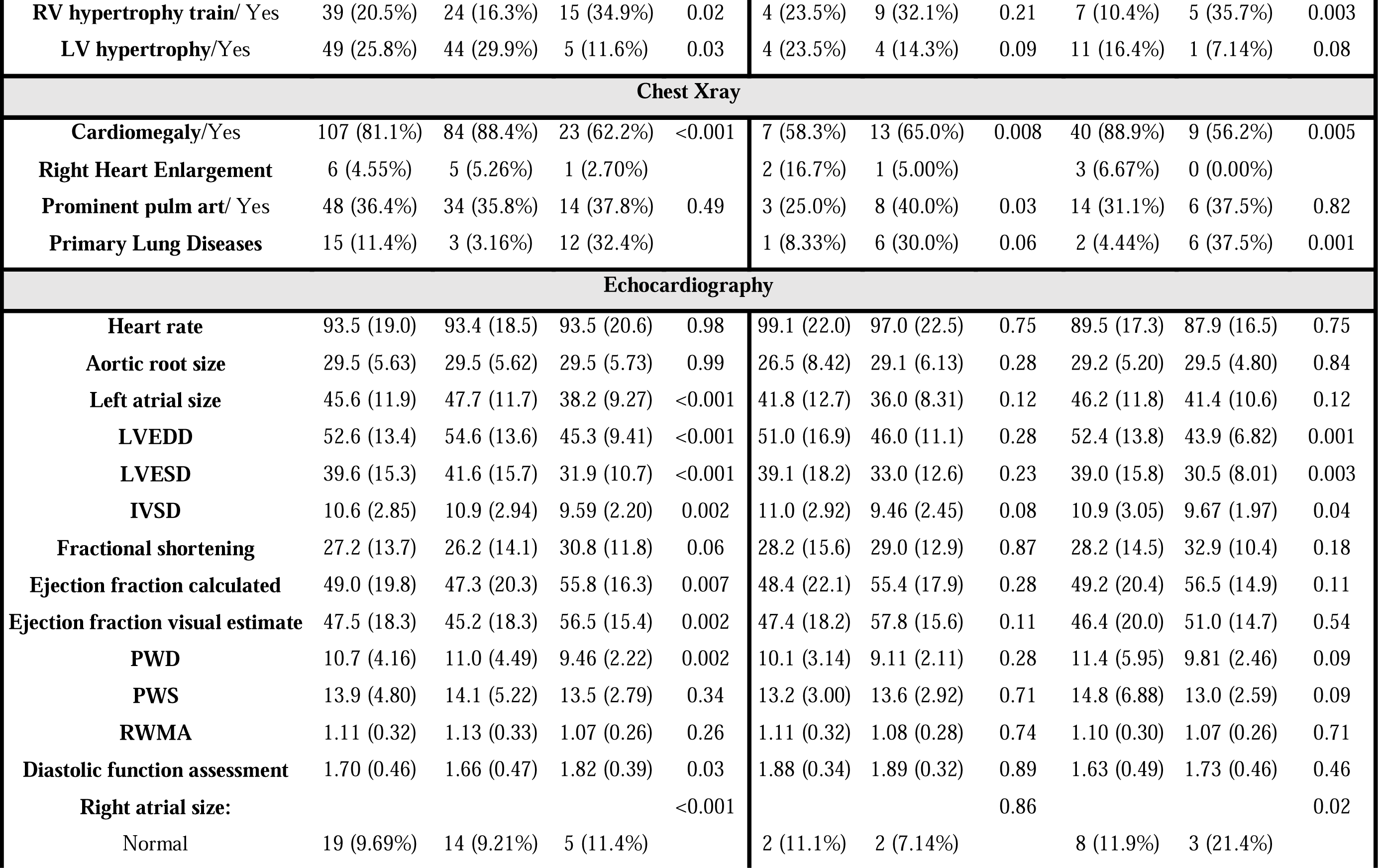
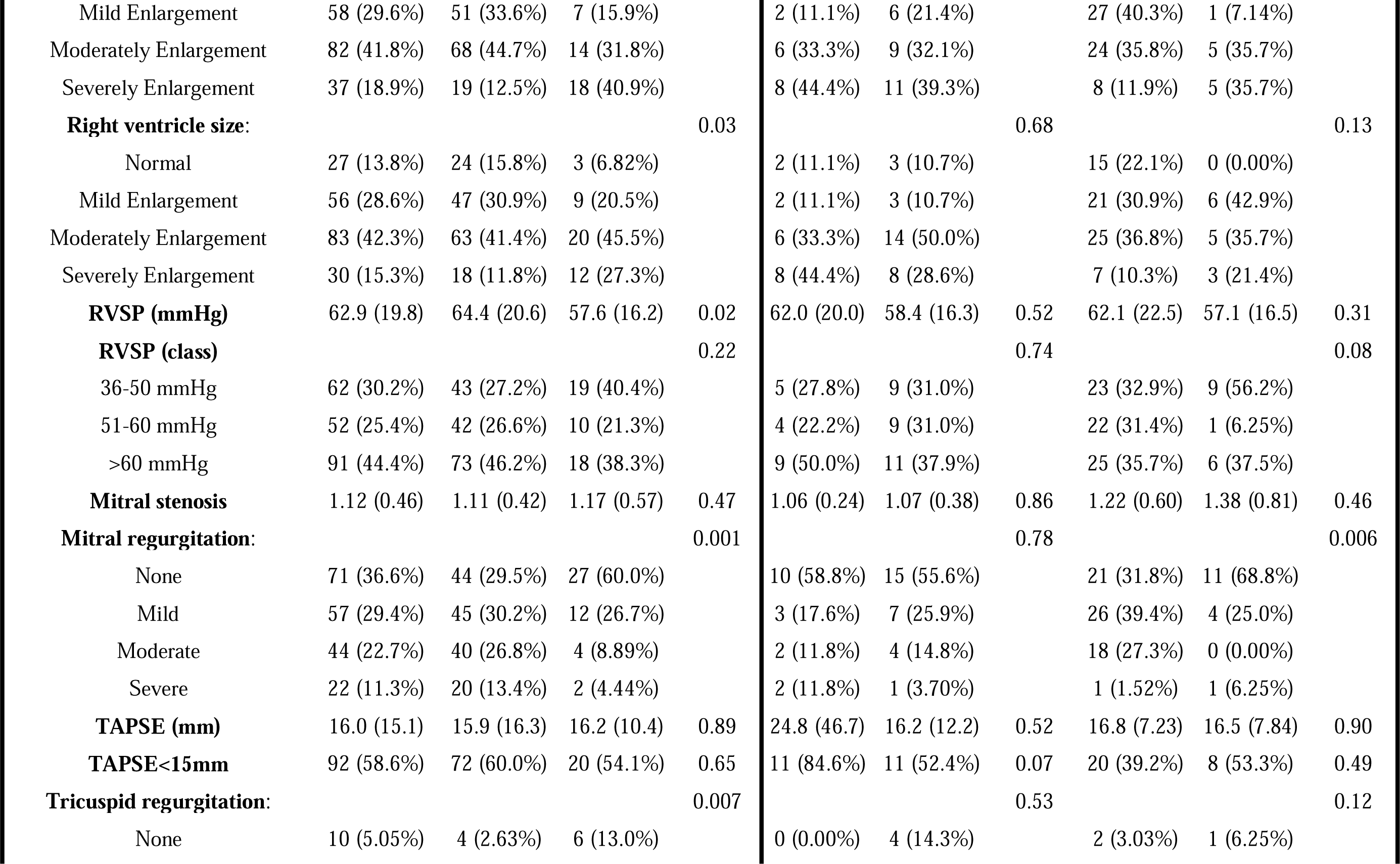
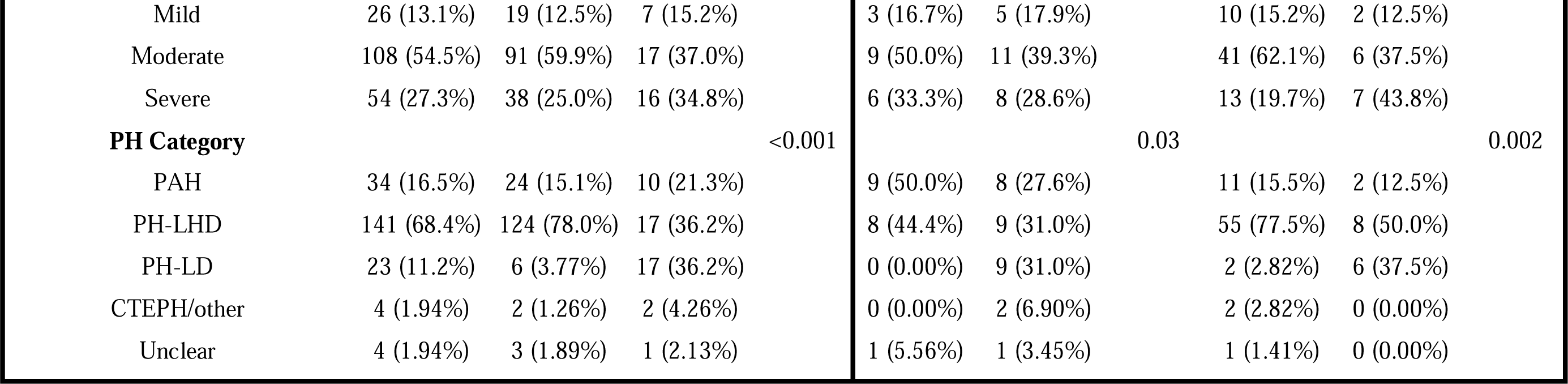
Electrocardiogram, X-ray, and echocardiographic findings of 206 patients included in the Pan African Pulmonary Hypertension Cohort by HIV-infection status and by history of tuberculosis.

### Characteristics of 134 participants with PH in the PAPUCO registry by HIV-status and TB (active or past)

Among the 206 PH patients included in the study, 134 (65%) had complete data on HIV-status. 47 (35%) of them were people living with HIV (PLWH, **Table 1**). 29 (62 %) of the 47 PH PLWH had HIV/TB coinfection, with 13 (45 percent) having more than one TB episode. In contrast, only 16 (18%)/87 PH patients who are HIV-uninfected had a history of TB. While not statistically significant, PH patients with HIV/TB coinfection were younger, unemployed, had a monthly income of less than $30, and lived in temporary shelters as compared to PLWH without a history of TB. They also tended to come from families with no history of CVD (21 vs 50 %), but to present with a history of other chronic lung diseases (35 vs 0 %) when compared to PLWH without a history of TB. During clinical examination, PH PLWH with a history of TB complained of fatigue less frequently (69 vs 100 %) than those without a history of TB or even those with no HIV/TB (69 vs 88 %). Furthermore, their BMI was lower (22 vs 28 kg/m^2^) than PLWH without a history of TB. Similarly, they tended to have lower oxygen saturation during pulse oximetry (94 vs 97 %), a higher frequency of raised JVP (90 vs 68 %), more normal cardiac auscultation (24 vs 0 %), and higher CRP (33 vs 11) when compared to PLWH without a history of TB. Patients with PH who had a history of HIV/TB coinfection had a lower chance of obtaining phosphodiesterase therapy than those who did not (6 vs 0 %).

**Table 2** summarizes the findings of the ECG, chest X-ray, and echocardiography. Atrial fibrillation was often observed on ECG in PH patients with TB, whether co-infected with HIV or not, when compared to those without a history of TB. However, when compared to PH patients without a history of TB, P-pulmonale was more common in PH patients coinfected with HIV/TB, but RV hypertrophic strain was more common in those who were HIV-uninfected. Cardiomegaly was more prevalent in patients with PH but no history of TB. We did find an exception in a subset of PH patients with HIV infection, where those with HIV/TB coinfection had a considerably higher proportion of cardiomegaly than those without such co-infection. Primary lung illnesses on X-ray were more common in PH patients without HIV but with a history of TB than PLWH. When compared by HIV status, echocardiographic findings revealed a significant difference between PH patients with a history of TB and those without such a history in terms of left ventricle circulation disorders, which were more common in patients without HIV infection and a history of TB. Regardless of HIV infection status, PH patients with a history of TB had a lower average RVSP than their peers with PH without a history of TB. Among PLWH, those with TB were usually classified as PH-LD or CTEPH, but those without a history of TB were mostly classified as PAH, PH-LHD, or PH with unclear and/or multifactorial mechanism (group 5).

### TB history as predictor of death in patients with PH

Figure 1 depicts the poor prognosis associated with recurrent TB in PH patients. **Table 3** provides models that account for age, gender, and RVSP in order to investigate the influence of TB history and its relationship with diabetes, HIV infection, and chronic lung illnesses in predicting death in PH patients. Firstly, we found that, while having a relatively minor clinical effect, every 1 mmHg increase in RVSP was strongly related to risk of death among patients with PH in all four models. Secondly, we found that patients with a history of TB were 1.82 times more likely to die from PH (aHR: 1.84; 95 percent CI: 1.00, 3.39; p=0.049) than those without TB history. Thirdly, when a history of TB coexisted with a history of other comorbidities known to worsen the course of TB, the hazards of death doubled or even tripled among individuals being followed for PH. The clinical effect observed did not reach the level of significance for diabetes history (aHR: 2.39; 95%CI: 0.32, 18.00; p=0.4), was borderline with HIV co-infection (aHR: 2.10; 95%CI: 0.97, 4.54; p=0.059) and was statistically significant for chronic lung diseases (aHR: 3.10; 95%CI: 1.47, 6.53; p=0.003).

**Figure 1:**
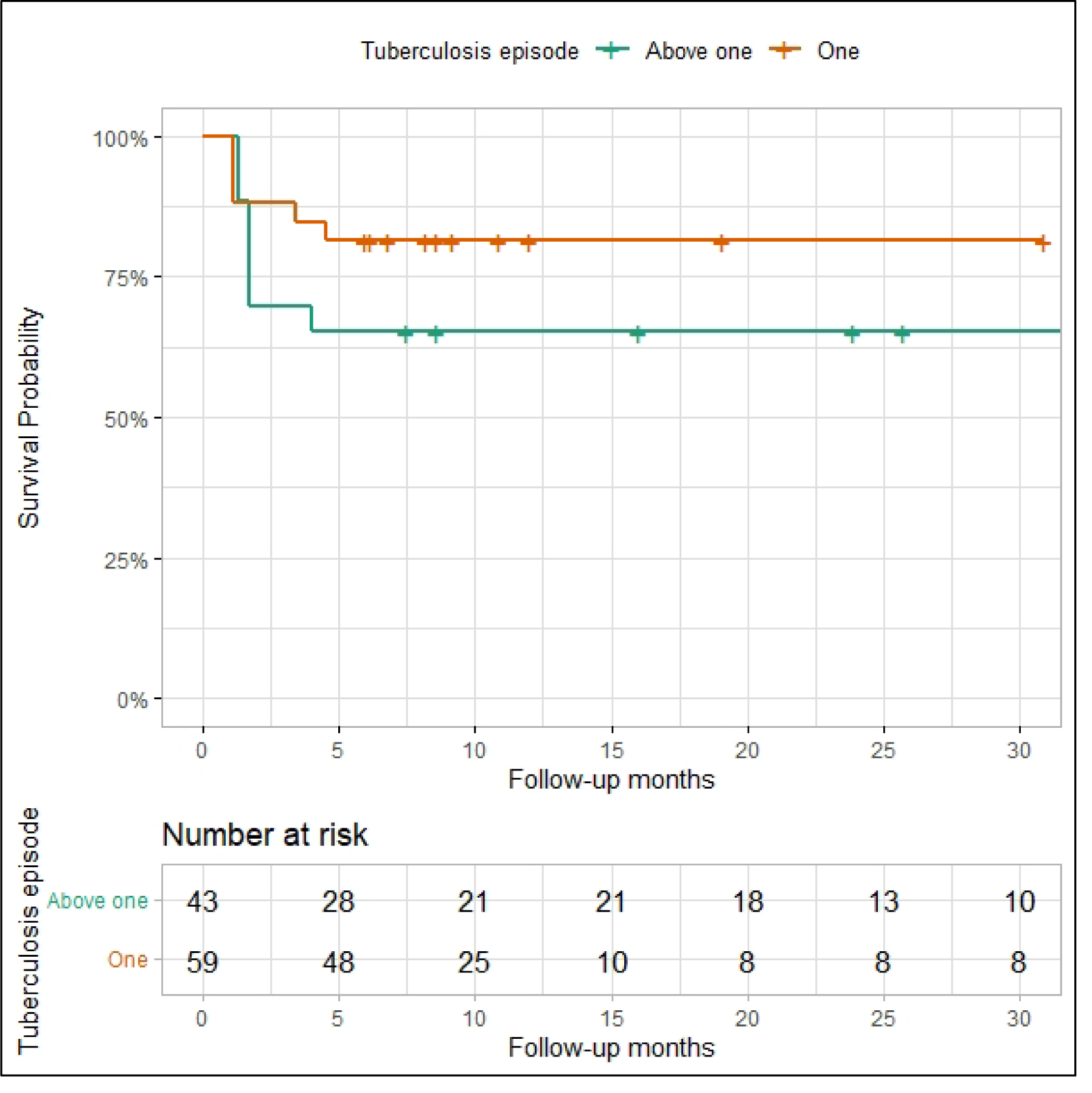
Kaplan Meier curve showing the effect of recurrence tuberculosis on the survival rate in 102 patients included in the Pan African Pulmonary Hypertension cohort. Note that three patients with missing data on TB recurrence were excluded from the current analysis.

**Table 3:** Tuberculosis and its interaction with diabetes mellitus, HIV-infection and chronic lung diseases in relation with hazards of death in 206 patients included in the Pan African Pulmonary Hypertension cohort. All the four models control for age and gender as well as for RSV used as measure of severity in the PAPUCO study.

| Variables | Model 1 |  |  |  | Model 2 |  |  |  | Model 3 |  |  |  | Model 4 |  |  |  |
| --- | --- | --- | --- | --- | --- | --- | --- | --- | --- | --- | --- | --- | --- | --- | --- | --- |
|  | aHR | 95%CI |  | P-value | aHR | 95%CI |  | P-value | aHR | 95%CI |  | P-value | aHR | 95%CI |  | P-value |
| Age | 1.01 | 0.99 | 1.02 | 0.23 | 1.01 | 0.99 | 1.02 | 0.24 | 1.02 | 1.00 | 1.04 | 0.08 | 1.01 | 1.00 | 1.03 | 0.14 |
| Sex | 1.09 | 0.64 | 1.85 | 0.74 | 1.08 | 0.63 | 1.85 | 0.77 | 0.80 | 0.42 | 1.51 | 0.49 | 1.13 | 0.66 | 1.93 | 0.66 |
| RVSP | 1.01 | 1.00 | 1.03 | 0.04 | 1.01 | 1.00 | 1.03 | 0.05 | 1.02 | 1.01 | 1.04 | 0.001 | 1.02 | 1.00 | 1.03 | 0.03 |
| Previous TB | 1.84 | 1.00 | 3.39 | 0.049 |  |  |  |  |  |  |  |  |  |  |  |  |
| Interaction |  |  |  |  | Previous TB # Diabetes |  |  |  | Previous TB # HIV Status |  |  |  | Previous TB # Chronic Lung Diseases |  |  |  |
| No-Yes |  |  |  |  | 1.13 | 0.39 | 3.27 | 0.82 | 1.94 | 0.77 | 4.92 | 0.16 | 0.41 | 0.06 | 3.07 | 0.39 |
| Yes-No |  |  |  |  | 1.88 | 1.00 | 3.54 | 0.049 | 1.68 | 0.59 | 4.73 | 0.33 | 1.32 | 0.59 | 2.96 | 0.50 |
| Yes-Yes |  |  |  |  | 2.39 | 0.32 | 18.00 | 0.40 | 2.10 | 0.97 | 4.54 | 0.059 | 3.10 | 1.47 | 6.53 | 0.003 |

Model 1 adjust for age, sex and RVSP; model 2 interaction between previous history of tuberculosis and diabetes; model 3 interaction between previous history of tuberculosis and HIV-infection; model 4 interaction between previous history of tuberculosis and history of Chronic lung diseases, other than tuberculosis. All models adjust for age, sex and RVSP. Under interaction term in variables (column 1), read: No-Yes: No previous TB but presence of associated comorbidity under exploration; Yes-No: history of previous TB but no history of associated comorbidity under exploration and Yes-Yes: presence of both history previous TB and of associated comorbidity under exploration. aHR: adjusted hazard ratio, CI: confidence interval, TB: tuberculosis

Please add a section on PH active TB and PH post-TB (see our systematic review)

## Discussion

### Main findings

In this secondary analysis study of the Pan African Pulmonary Hypertension Cohort (PAPUCO), a unique contemporary multi-country registry available to the continent, we assessed, for the first time, the effect of history of TB on the prevalence and hazards of death associated with PH in the region with important burden of HIV/TB coinfection while bearing in mind other emerging risk factors for TB such as diabetes and chronic lung diseases on rise on the continent as well. Firstly, we found that patients with history of TB were 1.82 time more likely of dying with PH than were PH patients without history of TB. Secondly, when history of TB was present concomitantly with history of diabetes, HIV-infection and chronic lung diseases, conditions known to exacerbate the course of TB, the hazards of death doubled or even tripled among patients with PH and was statistically significant for chronic lung diseases.

### Other important findings

Other main findings of our study include the fact that, approximately a quarter of patients with PH had history of TB. Patients with history of TB who developed PH were significantly younger (39 vs 53 years) and had lower socioeconomic status (SES), compared to patients without history of TB.

Further, we noted that patients with PH and with history of TB complained less for fatigue than those without history of TB and consequently performed well during six-minute walk test. They had reduced BMI, but an elevated CRP compared to PH patients without history of TB. Secondly, we found that, more than one third of those who had a confirmed test for HIV-infection were PLWH at the time of PH diagnosis and among them, more than half (62%) had current or previous TB. Further, we noted that primary lung diseases on X-ray were commonly reported in PH patients without HIV infection but with history of TB as compared to PLWH. Considering HIV infection status, echocardiography showed that left ventricle circulation disorders were predominant among patients without HIV infection and history of TB as compared to their counterparts with history of TB. However, regardless of HIV infection status, PH patients with history of TB had a less increased average of RVSP as compared to their counterparts who presented PH without history of TB. PH patients with history of TB had less chance of receiving phosphodiesterase treatment as compared to those without such history (6% vs 0%).

### Comparison with other studies

While our study is the first to assess the impact of TB in a cohort of patients being followed for PH, our findings are consistent with previous research on the burden and prognosis of PH in people with a TB history. Our findings included a percentage of people who are currently receiving TB treatment (28%). Right heart failure is not uncommon in patients with pulmonary TB, particularly in high-burden countries. In an observational study in India, 104 of 789 patients starting TB treatment were found to have PH; 73% of those had a previous history of TB. At the end of the study, mortality in patients with TB and PH was higher (22%) than in patients with pulmonary TB alone (9%), indicating a poor prognosis in those who developed PH secondary to TB (p<0.001)^7^. Furthermore, contrary to most national TB guidelines, it is now evident that TB care is far from ended once a microbiologically confirmed cure has been achieved, as TB survivors endure chronic disability and an increased risk of mortality, often referred to as post-TB disease and in particular post-TB lung disease (PTLD). Of the 122 million DALYs lost due to incident TB disease in 2019, 58 million (47%) DALYs were attributed to PTLD with estimates of per-case post-TB burden higher for younger people and in countries with high incidence rates^4^. Similarly, a recent meta-analysis focusing solely on post-treatment mortality among TB survivors (40 781 people and 6922 deaths) reported that, regardless of TB clinical outcome, people with TB were 2.9 to 3.76 times more likely to die than the general population or matched controls, with cardiovascular disease accounting for the majority of deaths (20%) ^2^.

Unfortunately, pulmonary vascular disease data in TB survivors is limited. In Sudan, a study of successfully treated TB patients who presented with shortness of breath documented the presence of PH in a cohort of symptomatic patients in a country with a high TB burden. Compared to patients with PH associated with chronic obstructive pulmonary disease (COPD), these patients were significantly younger (43 vs 66 years)^11^. Another study compared TB survivors who presented with PH and who had similar extent of fibrosis radiologically diagnosed. Regardless of exposure to tobacco smoking, they noted two distinct phenotypes in spirometry, COPD and restrictive changes as in diffuse parenchymal lung disease with the later phenotype resulting in a lower quality of life^19^.

Further, we noted that primary lung diseases on X-ray were commonly reported in PH patients without HIV-infection but with history of TB as compared to PLWH, which is most likely PTLD. It is unclear how the extent of the lesion affects the severity of PH in PTLD. PTLD on X-ray presents with fibro-cavitation and bronchiectasis and usually develops as a result of years of chronic TB, often multiple episodes of TB, treatment interruption, or drug resistant TB. PTLD is associated with high mortality rates and a field score of three was found to be an essential predictor of shorter survival indicating that PTLD has a poor prognosis, particularly in patients with more extensive lung destruction^20^. It is also not well documented how the extent of lesion might impact on the pathogenesis, clinical and prognosis of PH in TB survivors. In South Korea, a comparative analysis between patients with COPD and PAH was conducted. It appeared that patients in the PAH group (n=37) had a smaller lung volume (forced vital capacity percent predicted: 51.55 vs 72.37, p<0.001) and more extensively destroyed lungs (3.27 vs 2 lobes, p<0.001) than the non-PAH group (n=16). As a result, PAH was associated with the severity of lung destruction in PTLD, but the risk of exacerbation and mortality did not differ significantly between patients with and without PAH^21^.

### Impact on the policy and current practice

Over the last decade, there has been a surge in interest in PTLD^22^. However, three major issues remain patent as of today. First, TB as a disease of poverty has seen less interest toward directing research and policy on addressing associated pulmonary vascular and cardiovascular diseases as the main cause of death after successful treatment. To address the global disparities in TB follow-up care as well as global PH quality of care, scientific communities should consider testing and implementing improved, less expensive, and less invasive diagnostic methods^10^. Further, despite the negative impact demonstrated on quality of life after TB treatment, the effect of socioeconomic status (SES) on the disease process is not taken into account in current guidelines and management algorithms^23^. Consequently, patients are referred to care facility at the late stage when presenting with worsening dyspnea and are incorrectly treated for relapse of TB or are prescribed inhaled bronchodilators without complete lung function assessment including diffusing capacity of the lungs for carbon monoxide (DLCO) or documentation of airflow obstruction. This mismanagement can be avoided by implementing clear algorithms for earlier diagnosis and referral system of both TB and PTLD at primary care facility^24^. For example, while controversial, radiologically, patients with PH who have a history of TB have a poor but negative co-relationship with the degree of fibrosis^19^. Although clinicians can easily diagnose PTLD in patients with destroyed lungs on X-ray and with a history of TB, they can also predict the outcome in patients with extensive lung destruction^20^. At ECG, P-pulmonale, qR pattern in V1, and heart rate were found to be associated with prognostic factors in severe PH and may be a useful tool in follow-up^25^. We noted that although at a very small clinical effect, every 1 mmHg increase in RVSP was significantly associated with the hazard of death among patients with PH in all four models. It therefore suggests that in settings with limited resources, training for accurately assessing RVSP using echocardiography assist decision making for PH management.

Second, the definition of PTLD for clinical practice and associated risk factors has to be refined and endorsed by different societies and task forces, and the biological mechanism leading to pulmonary vascular disease needs to be elucidated. The clinical phenotype of PTLD is mosaic and includes cavitation, bronchiectasis, fibrosis, and other advanced structural changes such as lung distortion. While the precise mechanisms are not yet fully understood, four important components (granuloma formation and resolution process, cytokine production effect, transcription factors effect, and enzymes such as matrix metalloproteinases) have been postulated. In addition, host, pathogen and environmental factors are also projected to be key for such clinical and pathological presentation^3^. As complex as this might sound, user-friendly guidelines based on simple clinical and paraclinical factors can assist busy clinicians for risk factor screening for both PTLD and PH, allowing for proper resource allocation especially in LMICs. For example, in a previous population-based cross-sectional study of 441 randomly selected PTB survivors living in 13 remote health zones with high TB burden in the Democratic Republic of the Congo, we found a high prevalence of chronic cough, with the odds of having it doubled with exposure to household air pollution from biomass used for domestic energy^26^. Furthermore, various co-morbidities that aggravate the course of TB, such as previous COPD, diabetes, and HIV co-infection, may have a role in the pathogenesis and severity of PTLD and hence raise the red flag. While we found a significant frequency of HIV/TB co-infection, the effect of HIV during TB co-infection on PH survival was marginal. Previous research, while preliminary and limited, has revealed that HIV co-infection may be associated with less severe PTLD^3^.

Third, we found that patients with TB history were more likely to be classified in group 3, PH-LD (36 vs 4%), compared to their counterparts. The critical question remains whether TB-associated PH should be considered group 1 PAH such as HIV-associated PH or schistosomiasis-associated PH, group 3 PH-LD or group 5 with unclear and/or multifactorial mechanism. This has large implications on treatment options, as treatment is available for group 1 with very little options for group 3. However, most group 1 treatment options are either not available in LMICs or prohibitively expensive. A complicating factor is the common HIV/TB co-infection in LMICs, and multifactorial causes of PH are most likely. Hence, a clear definition and staging is warranted in order to develop treatment for TB-associated PH in the context of HIV infection. On one hand, researchers from high income countries barely address the interplay role between TB and PH and the most recent ESC/ERS guideline^8^, only evoked TB as a potential etiology of fibrosis mediastinitis in PH with unclear etiology. On the other hand, researchers from high TB burden countries have independently highlighted this divide following their clinical practice. In a commentary, Verma^24^ stated that in high-income countries, pulmonary TB is rarely cited as a cause of the development of PH. However, in India, a high burden country, it is common that patients who have been treated for pulmonary TB present with features of right heart failure. Allwood et al.^9^ acknowledged that there may be a discrepancy between what is seen in clinical reality and what is reported on post-TB pulmonary vascular disease and PH. They speculated that the difference could be explained by the lower TB incidence and, as a result, the lower prevalence of TB evolving to advanced lung destruction in countries where PH is being studied compared to what practitioners saw daily in high TB burden regions. If *de novo* guidelines cannot be developed, we support the need for adapting current PH and TB guidelines to include TB/PTLD and PH respectively.

### Strengths and limitations

The main strengths of our study includes the assessment for the first time of the effect of previous TB on the prognosis in a cohort of PH patients, as well as using robust multi-country and multicenter data from PAPUCO study. We recognize the distinction between transthoracic ultrasound and the gold standard of right catheterization in the diagnosis and management of PH. We also believe that, due to the scarcity of qualified human resources and equipment across sub-Saharan region, as well as the significant risk factors for pulmonary vascular and cardiovascular diseases and PH in young people living in the region, a simple algorithm involving RVSP can provide data on PH and guide practice where it is most needed. Further, the younger age of our participants and a significant proportion of PLWH and TB may restrict the generalizability of our findings to non-similar settings. This is an excellent opportunity for multi-disciplinary and geographically matched research involving infectious disease specialists, cardiologists, and pulmonologists to strengthen TB control activities (prevention, diagnosis, and treatment) while the effort to anchor PTLD care across the TB care continuum is ongoing. The research might echoed previous and emerging questions on the topic such as the strength of the association between pulmonary TB and PH across the spectrum of parenchymal abnormality in TB patients, the poor correlation between the degree of right heart failure and the degree of radiological changes, the relationship between the degree of right heart failure and the degree of airway obstruction/restriction, regardless of radiological changes, the time for PH commencement during and after TB treatment as well as the role of other risk factors like tobacco smoking, exposure to air pollution, drug use, diabetes or HIV-infection or COVID-19 co-infection in the development of PH^9,27^.

## Conclusion

In this cohort study of African patients followed for pulmonary hypertension, TB history and HIV-infection were found to be highly prevalent and associated with poor survival outcomes. Our findings represent a call to address the burden of pulmonary hypertension associated with TB during and after treatment, which affects a disproportionate number of working-class people from low-income communities.

## Funding

Financial support was provided by unconditional research grants from the Pulmonary Vascular Research Institute (PVRI) [Grant number 422348] and Bayer Healthcare Berlin [Grant number 411278]. The General Medicine & Global Health (GMGH) Research Unit of the University of Cape Town provided institutional support. PDMCK is supported by the NIH/Fogarty 1D43TW010937-01A1 (the University of Pittsburgh HIV Comorbidities Research Training Program in South Africa— Pitt-HRTP-SA). SLM, KW, and FT are funded by the European Union (grant number RIA2017T-2004-StatinTB)

## Author contributions

Conceptualization, P.K., M.S., K.K., A.D., A.M., A.D., K.S. and F.T.; methodology, P.K., M.S., K.K., A.D., A.M., A.D., K.S. and F.T.; software, P.K., S.M., K.W., I.M. and F.T.; validation, P.K., M.S., K.K., A.D., A.M., A.D., K.S. and F.T. formal analysis, P.K., investigation, I.M., S.M., KW., M.S., K.K., A.D., A.M., A.D., K.S. and F.T. resources, K.S., A.M. and F.T.; data curation, P.K., I.M., S.M., K.W., M.S., K.K., A.D., A.M., A.D., K.S. and F.T. writing—original draft preparation, P.K.; writing—review and editing, I.M., S.M., K.W., M.S., K.K., A.D., A.M., A.D., K.S. and F.T.; visualization, P.K. and F.T.; supervision, K.S. and F.T.; project administration, P.K. and S.M.; funding acquisition, K.S. and F.T. All authors have read and agreed to the published version of the manuscript.

## Data Availability

All data produced in the present study are available upon reasonable request to the authors

## Acknowledgement

The authors thank the PAPUCO study teams at all African sites for their help with collecting medical reports and retention of study patients. This publication was produced by members of the StatinTB consortium which is part of the EDCTP2 programme supported by the European Union (grant number RIA2017T-2004-StatinTB).

## Data availability

Data is available from the corresponding author upon a reasonable request.

## Conflict of interest

We report no conflict of interest.

## Notes

### Competing Interest Statement

The authors have declared no competing interest.

### Author Declarations

South Africa (University of Cape Town faculty of health science: FWA0000163)

